# A more complete picture: Capturing single nucleotide variant diversity in extended-spectrum beta-lactamase producing *Escherichia coli* using post-enrichment metagenomics

**DOI:** 10.1101/2025.11.12.25340063

**Authors:** Sarah Gallichan, Tommi Mäklin, Esther Picton-Barlow, Claudia McKeown, Sally Forrest, Jukka Corander, Maria Moore, Nicholas A Feasey, Eva Heinz, Fabrice E Graf, Joseph M Lewis

**Author notes:** **Corresponding author:** Sarah Gallichan Joseph Lewis.

## Abstract

Inferring transmission relies on accurately distinguishing between isolates from the same source and those from different sources, and high-quality genomic data is frequently used to model transmission scenarios. The post-enrichment metagenome sequencing (pe-MGS) method uses a sequencing approach to analyse the diversity of a target pathogen enriched by pre-culturing, and has been effectively used to analyse the transmission of nosocomial infections. However, a direct comparison of single nucleotide variant (SNV) call accuracy, cost and feasibility between single colony whole genome sequence (sc-WGS) data and pe-MGS for an antimicrobial resistant bacteria of clinical importance, extended-spectrum beta-lactamase producing *E. coli* (ESBL-EC), is required for implementation in large-scale clinical studies. A spiked stool sample and rectal swabs from six study participants were pre-enriched in buffered peptone water and cultured on MacConkey agar with 1mg/L cefotaxime. Seven single colonies were picked, and the remaining biomass of all colonies was collected from each plate, sequenced and analysed using the mSWEEP/ mGEMS pipeline. We created a custom SNV calling workflow that allows heterozygous SNVs in a bacterial population, and found that the choice of reference changed the number of measurable SNV distances between the sc-WGS and pe-MGS. Using our custom workflow with a core-gene reference captured 99% of all the SNV calls from multiple sc-WGS data in the pe-MGS data of the same culture. The plate sweep method offers a feasible, cost-effective alternative to multiple single colony picks for describing within-host ESBL-EC diversity. The workflow we developed allows for effective SNV calling from pe-MGS data that was comparable to SNV calls from multiple sc-WGS data from the same sample.

**Abbreviations:** AMR, antimicrobial resistance; ESBL-EC, extended-spectrum beta-lactamase producing *Escherichia coli*; ST, sequence type; SNV, single nucleotide variant; BPW, buffered peptone water; sc-WGS, single-colony whole-genome sequencing; pe-MGS, post-enrichment metagenome sequencing

**Impact statement:** For bacterial species with high within patient diversity and within-genome variation such as the opportunistic pathogen *E. coli*, capturing the full diversity is essential to identify transmission events. Pre-enriching the species of interest from patient samples, and then sequencing all colonies, post-enrichment metagenomics (pe-MGS) promises to be a cost-effective, efficient method capturing the full diversity. To estimate transmission events with high confidence and make it applicable for hospital transmission studies, single-nucleotide variants (SNVs) have to be identified with an equivalent resolution as would be achieved when using single-colony whole-genome sequencing on all colonies. Here we present a proof-of-concept study on a set of stool samples and rectal swabs from healthy participants, where we developed a new workflow tested against these control samples. All samples were analysed using both single-colony WGS (sc-WGS) and pe-MGS from the same plate, following pre-enrichment for the species and phenotype of interest (drug resistant *Escherichia coli*). This direct comparison allowed us to assess the reconstruction of SNVs between the two approaches on clinically relevant sample types at the highest resolution. We show that by using a newly developed SNV calling workflow, a core-gene reference allowed us to identify comparable SNVs in the pe-MGS to the sc-WGS sequence data. The pe-MGS offers a cheaper and time-saving alternative to multiple sc-WGS and is thus more feasible to integrate into public health settings for routine surveillance. This would provide a robust basis to significantly improve our detection of transmission events and thus understanding of transmission routes, and allow for the targeted implementation of preventative measures.

**Data summary:** The short-read sequence data generated in this study have been submitted to the European Nucleotide Archive (ENA, https://www.ebi.ac.uk/ena/browser/view/PRJEB101999), and their individual accession numbers are listed in **Supplementary Table 1**. Github repository: https://github.com/joelewis101/TRACS-liverpool/tree/main/bioinformatics. All protocols developed and utilized in this study have been detailed and provided in the article and supplementary data files.

## Introduction

Identifying bacterial pathogen transmission events is important to understand spread, highlight those at risk of infection and implement targeted measures to prevent further infection^1^. In order to infer pathogen transmission, it is necessary to identify the presence or absence of differences between isolates, a process traditionally called “typing”. Typing methods rely on accurately and unambiguously distinguishing between isolates from the same source and those from different sources^2^. Genomic data provides a high-resolution method to type pathogens by measuring the genetic distance of pathogens at single nucleotide variant (SNV) resolution. This requires sequence data with sufficient read quality and depth to discern between highly similar strains^3,4^. Given the still substantial cost of sample preparation and WGS at the necessary read depth and quality, transmission studies often rely on sequencing a single bacterial colony from each sample which gives high resolution but no information of the diversity^5–7^.

For pathogens that exhibit largely clonal outbreaks and are not also part of the healthy human microbiome, such as *Salmonella enterica* serovar Typhi, single colony whole genome sequencing (sc-WGS) is often sufficient to capture transmission events^8^. However, for highly diverse opportunistic pathogens that are also part of a healthy microbiome, such as *Escherichia coli*, the chance of capturing transmission events with a single colony from complex microbial samples becomes more unlikely as there is substantial within-sample diversity, within which the strain of interest has to be identified. Studies of human stool samples suggest that while the analysis of five *E. coli* colonies from a sample can detect the dominant genotype at >99% probability, as many as 28 single colonies per stool sample are required to detect minor genotypes (at 90% probability)^9,10^. However, sequencing and analysis of multiple single colony picks to cover the diversity and detect low-abundance strains is time consuming, expensive, and not practical in routine transmission studies. For *E. coli*, a bacterium that is spread by the faecal-oral route where gut colonisation generally precedes infection, and which is used as a marker of bacterial transmission^11^ (i.e. by Water Sanitation and Hygiene [WASH] specialists), pragmatic approaches to describing full *E. coli* diversity within stool samples, at scale, are needed.

Metagenomic approaches offer a potential alternative to capture the full microbial diversity within a complex sample like stool^12^. While shotgun-metagenomics on direct DNA extractions from clinical samples has been used in clinical settings to identify pathogens, it may have limited success, or require expensive ultra-deep sequencing, to detect bacteria present at low abundances in complex microbial communities. This is often the case with *E. coli* in human stool samples, which generally are in abundance of the order of 1% within the human gut microbiome^13,14^. Thus, addressing the within-species diversity of *E. coli* in stool using shotgun-metagenomic sequencing is currently not feasible without sequencing depths that are prohibitively expensive. To strike a balance between the limited single colony analysis and the full microbial diversity of a shotgun-metagenomic approach, a variety of novel approaches, variously termed plate-sweep metagenomics, limited diversity metagenomics and post-enrichment metagenomics have been proposed to resolve these issues. For the purposes of this manuscript we will use the term post-enrichment metagenomics (pe-MGS).

These approaches typically start with a selective enrichment step to enrich for the bacteria of interest (for example *E. coli*) in broth, or an agar plate or both, overcoming the challenge of the low abundance of the organism of interest in raw stool. It proceeds by using all grown colonies (e.g. plate sweep, the picking of all colonies from a selective culture plate) for one combined DNA extraction and a single metagenomics sequencing run, and subsequent *in silico* reconstruction of the unique strains present in the sample^15^. The advantage of this strategy is the removal of DNA from host or bacteria not of interest, which are in a high proportion in stool samples, allowing for specific focus on the species of interest or drug-resistant phenotype, and the potential for a full description of bacterial diversity at much reduced labour and sequencing cost. The plate sweep method has been effectively used to analyse the within-sample diversity of several pathogens^16,17^, including the transmission of community-acquired *Streptococcus pneumoniae* as well as nosocomial infections *Klebsiella pnemoniae, E. coli,* Enterococcus species*, Pseudomonas aeruginosa,* and *Staphylococcus aureus*, in a clinical setting^18–^20_._

In this proof-of concept study, we assessed the feasibility of describing diversity of extended-spectrum beta-lactamase producing *E. coli* (ESBL-EC) in stool samples using pe-MGS and the mSWEEP/mGEMS^15^ workflow. ESBL-EC are resistant to aminopenicillins and third-generation cephalosporin antibiotics, which are commonly used as the first-line treatment for Gram-negative bacterial infections. Infections with ESBL-EC result in higher morbidity and mortality, and longer hospitals stays^21–23^. Here we compare the pe-MGS to the gold standard of sc-WGS to detect differences between highly related ESBL-EC strains in order to inform a healthcare associated ESBL-EC transmission study. We compare the predicted ESBL-EC diversity from human stool and rectal swabs based on the SNV call accuracy, cost, and feasibility, between these approaches.

## Methods

### Sample preparation

Stool from three healthy adult volunteers (individuals not on antibiotics or experiencing any gastrointestinal problems) was mixed and a slurry created for use as a model stool sample (LSTM Research Tissue Bank RTB/2022/007). A spiked stool sample was created with of two clinical ESBL-EC strains (a ST167 and ST131, previously isolated from human stool samples in Malawi^17^) mixed at equal ratio at 10^4^ CFU/mL, 10 μL spiked into 1 mL of pre-heated stool slurry (37 °C) and vortexed thoroughly. Rectal swabs which were collected from adult study participants (≥ 18 years) residing in hospitals and care homes in Liverpool as part of the TRACS-Liverpool study (TRACS: 22/NW/0343)^24^. The spiked stool and rectal swabs were then cultured according to an optimised method to recover ESBL-EC^25^. Briefly, stool and rectal swabs were pre-enriched for 4 hours in Buffered Peptone Water (BPW) at 37 °C while shaking at 220 rpm, 10 μL of enriched culture was then spread on cefotaxime (1 mg/ L) supplemented MacConkey agar and incubated at 37 °C overnight. Seven single colonies were picked using a 1 μL inoculation loop, spread on separate cefotaxime supplemented MacConkey agar plates (single colony picks) and incubated at 37 °C overnight. The remaining biomass was then scraped off and placed into a 1.5 mL tube (plate sweep), resuspended in PBS and extracted using the MasterPure Complete DNA and RNA Purification Kit (Lucigen, Wisconsin, USA) according to the manufacturer’s reccomendations. The same was done for the extraction of the single colony derived plates the following day.

### Sequencing

The DNA concentration and integrity were measured using the Qubit^26^ and the TapeStation System^27^, respectively. The extracted DNA from the single colony plates and the plate sweeps was then sent for Illumina sequencing, at depths of 1 Gb and 6 Gb respectively, to Azenta Life Sciences (Frankfurt, Germany). Data were obtained as paired-end 150 base pair reads. Adaptor sequences were removed using fastp version 0.23.4^28^ and the quality of the trimmed reads was assessed using the quality metrics provided by FastQC version 0.11.9^29^.

### Bioinformatic analysis

Reconstruction of bacterial strains within the sc-WGS and pe-MGS was done using the mSWEEP (v 2.2.0) and mGEMS (v 1.3.3) algorithms ^15,16^, using a curated reference database from a previous study^30,31^ (available from Zenodo^32^) consisting of 14,438 *E. coli* assemblies. The sample reads were first pseudoaligned to the reference database using Themisto (v 3.2.2)^33^, then mSWEEP was used to calculate the likelihood and relative abundance estimates of each reference PopPUNK (version 2.6.3)^31^ cluster being in the sample using the pseudoalignment results. The sample reads were then binned by running the mGEMS algorithm from mSWEEP with the –bin-reads option. The mGEMS read bins for each strain were quality controlled using the demix_check tool (v 1.0)^34^ and scored (1 – highest confidence, 4 – lowest confidence) based on the mash (v 2.0)^35^ distance between the binned reads and the sequences in the reference database. These samples were then extracted from the relevant read bins using mGEMS for use in SNV analysis. Multi-locus sequence typing was done using ARIBA (v2.11.1)^36,37^ with the seven-gene Achtman scheme^38^.

### Reference genomes

A closely related, Phylogroup B2, ST131 *E. coli* complete genome (Accession number: NC_000913, plasmids removed); a distantly related, Phylogroup F, ST1485 *E. coli* complete genome (Accession number: NZ_CP042896, plasmids removed); and a *E. coli* core-gene list were used as references. To create an *E. coli* core-gene list for use as a reference, the genes present in 99% of a pan-genome from a previously curated collection of 10,000 *E. coli* genomes^39^ were extracted from the pan-genome using SeqKit version 2.8.2^40^, resulting in 2 384 genes, with a total length of 2.29 Mb.

### SNV calling workflow

The sc-WGS were mapped to references and the SNVs were called using snippy version 4.3.6 (https://github.com/tseemann/snippy). A core genome alignment of all the single pick sequences was created using snippy-core (with default settings). All non-nucleotide characters, such as ‘n’, were removed from the multisequence alignment using the snippy-clean_full_aln and the resulting SNVs were extracted using snp-sites (with the -c option to output exclusively columns containing ATCG). A pairwise SNV distance comparison between each of the single picks in the sample was then created using snp-dists version 0.8.2 (https://github.com/tseemann/snp-dists). To account for the possibility of heterozygous SNVs present in mixed samples in the sweep samples, we modified the settings of snippy for these samples as follows: the SNVs were called using freebayes version 1.3.6^41^ (using the --pooled-continuous option) from the binary alignment map (BAM) file as produced by snippy using bwa-mem. The raw variant call files were filtered to include all variant types with a phred-scored quality of more than 100 (QUAL > 100) a depth of at least 20 (DP >= 20) and supported by at least 85% of reads (A0/DP > 0.85) at a given site using bcftools version 1.20^42^. In addition a sliding window approach was used to mask regions that had clusters of SNV calls, which could be sites of recombination or bacteriophage inserts that would influence the accuracy of transmission inference. The sliding window worked according to an algorithm^37^ whereby the rate of SNVs within an alignment was calculated within a set window (window size = length of reference genome/ total number of SNVs) and if a significant number of SNVs (p = 0.05) were captured in the window then those SNVs were excluded. Sites with low quality SNV calls (QUAL < 100), and/or had low sequencing depth (DP < 20) were masked across all SNV calls when comparing SNP distances. Heterozygous variant types (GT not equal to 1/1) were masked in sc-WGS. This SNV calling process was condensed into a Snakemake (version 8.29.2)^43^ workflow that is accessible from the TRACS-Liverpool Github page (https://github.com/joelewis101/TRACS-liverpool/tree/main/bioinformatics/snakemake).

### Sequence depth

To identify the optimum, we simulated different sequencing depths by subsampling reads; the number of reads that approximated 10x, 15x, 20x, 25x, 30x, 100x and 500x sequence depth **(Supplementary Table 2)** were randomly subsampled using Seqtk (version 1.4.)^44^ from the plate sweep sequence data. The subsampled sequence data was then analysed through the mSWEEP/ mGEMS pipline and the custom SNV calling workflow using the core *E. coli* gene list as a reference. To estimate the plate sweep sequence depth that can be used to accurately capture 99% of the combined single pick SNV calls, a logistic regression model was fitted to the rarified plate sweep sequence data using the nlstools R package version 2.1-0^45^.

## Results

### Calibrating a workflow to analyse SNV diversity in plate sweeps

Since tools to identify SNVs in bacteria are designed for homozygous samples and not to identify variants in mixed samples, heterozygous SNVs as present in the plate sweep sequence data were considered errors by default by tool expecting input from a homozygous population as standard for bacteria WGS. Therefore, we developed a custom SNV calling workflow that allows calling the heterozygous SNVs in sweep sequence data and directly compares the SNV calls in the sweep data to the pooled single pick data **(Figure 1)**.

**Figure 1.**
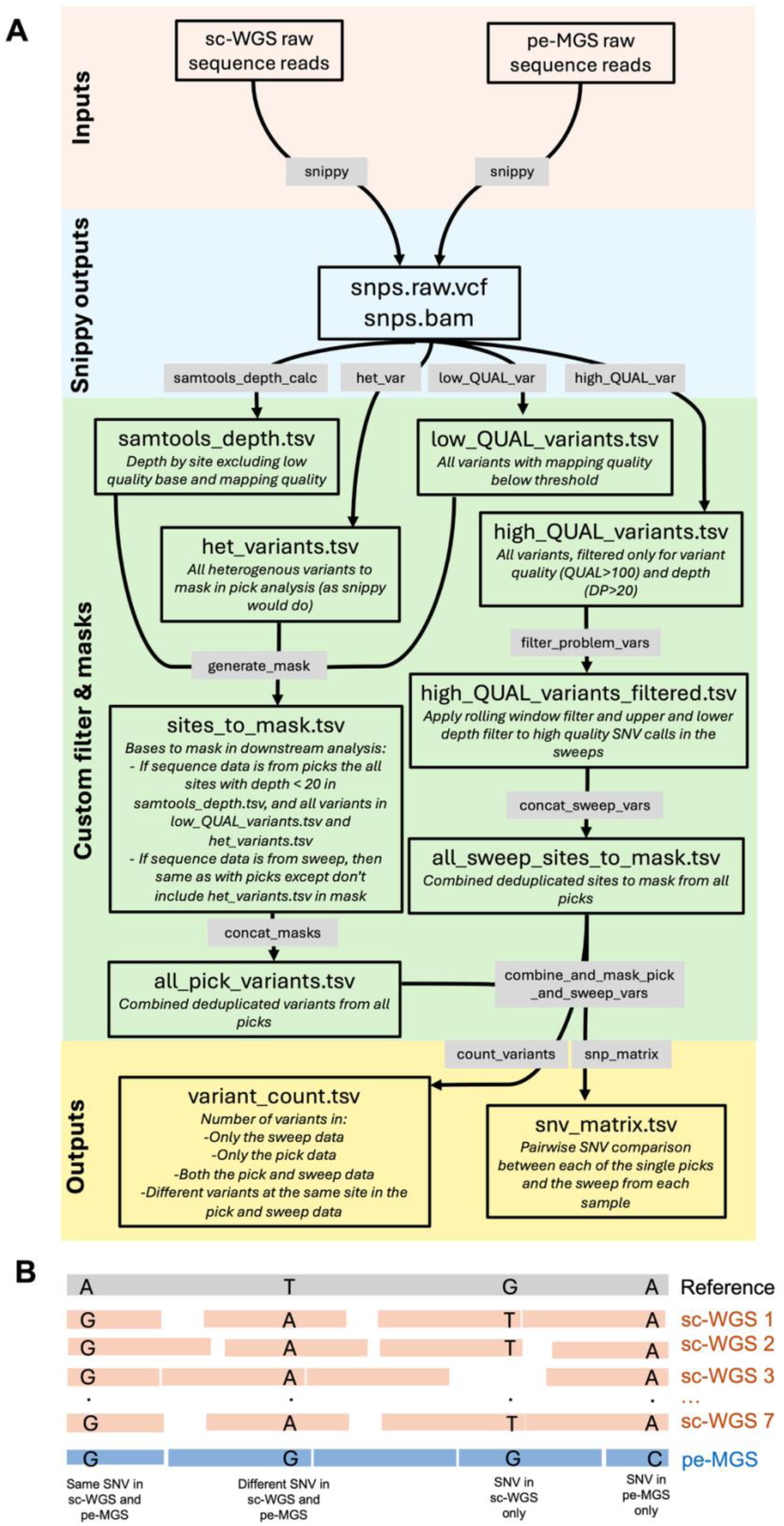
**A)** Overview of the custom SNV calling workflow that directly compares the SNV calls in the sequence data of multiple sc-WGS and a pe-MGS from the same sample. Grey boxes correspond with rules in the Snakemake workflow. **B)** Graphic illustration of the comparison of sequence reads (colour blocks) from multiple sc-WGS and a pe-MGS from the same sample. The black letters indicate SNVs in each of the sequence types and the descriptions below indicate how the SNV comparisons were grouped in the final variant count output from the workflow.

To calibrate this custom workflow, we used sc-WGS and sweep sequence data from a spiked stool sample. We added a mix of two well characterised clinical ESBL-EC strains (ST167 and ST131) that we assumed were clonal and would result in no measurable SNV distances between sc-WGS and pe-MGS data for each strain. We then directly compared the SNV calls using the Snippy tool and our custom SNV calling workflow in the sc-WGS and corresponding pe-MGS data from each of the two ESBL-EC strain spikes using three different references (a core-gene reference and two complete genome references, *E. coli* ST131 and ST1485) **(Figure 2)**. Overall, the SNV calls differed according to the reference used, with the highest number of SNVs calls when the ST1485 reference was used in both the custom SNV calling workflow and the Snippy tool **(Figure 2)**. Only the custom SNV calling workflow with the core gene reference produced SNV matrices with no measurable SNV distances between sc-WGS and pe-MGS from the spiked stool as we expected **(Figure 2)**.

**Figure 2.**
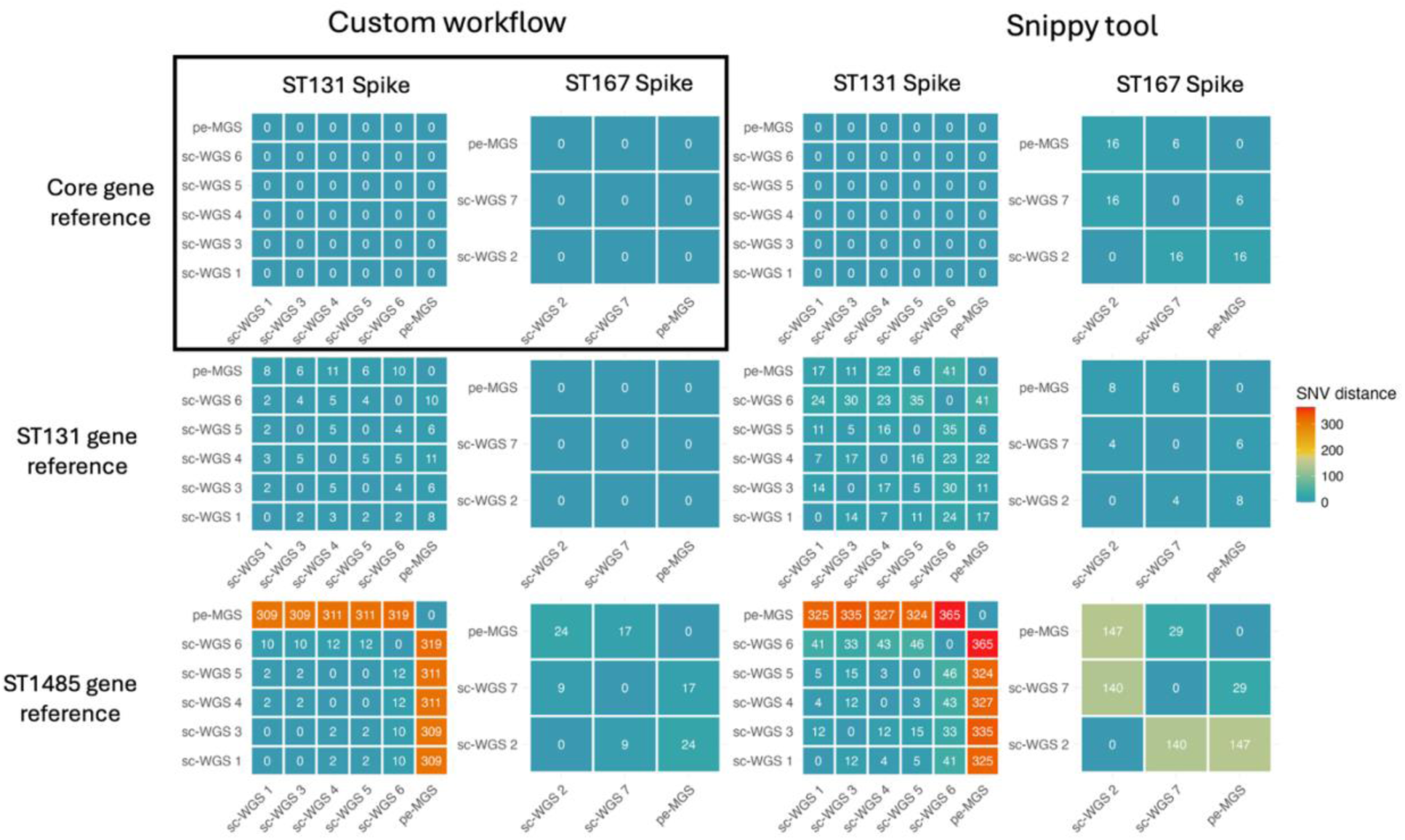
SNV distance matrices for the two ESBL-EC STs (five single colonies were ST131 and two single colonies were ST167 with corresponding sweep reads for each strain) in the spiked stool sample comparing snippy and our custom SNV calling workflow with three different references (Core-genes, ST131 and ST1485). The grey block highlights the workflow and reference that resulted in no measurable SNV distances as expected.

Since we assumed that the two ESBL-EC strains spiked into the stool were each clonal, we investigated why our custom workflow with the whole genome references (ST131 and ST1485 references) resulted in variation in the sc-WGS and pe-MGS data. The SNV calls were high quality, with sufficient read depth, and not masked by the sliding window filter **(Supplementary table 3)**. Thus, we assessed whether the variance was due to complex genomic regions such as hypervariable or repeat regions **(Supplementary table 3)**. The SNV calls mapped to the ST131 reference (n=17) were randomly spread across the genome and the majority of the calls, except one variant that mapped to an inner membrane protein region, did not have any associated protein data. However, the SNV calls mapped to the ST1485 reference (n=364) were distinctly clustered in hypervariable regions, such as genes encoding adhesin and membrane proteins ^46^, with 319 of the SNVs clustered to only 27 genes **(Supplementary table 3)**. To eliminate the variance due to hypervariability and the impact of different references based on the input sample going forward, we used the core gene reference for the analysis of clinical samples.

### Plate sweeps can be used to accurately capture SNV diversity in clinical samples

Rectal swabs from six TRACS-Liverpool study participants were pre-enriched, and selected for ESBL-EC on cefotaxime supplemented MacConkey agar. The resulting single colonies were submitted for sc-WGS, and remaining biomass for pe-MGS. Seven colonies were picked for each study participant where possible for sc-WGS; two participants (1 and 3) only had 6 and 4 visible colonies on the selective agar, respectively, which we thus selected. Sc-WGS indicated that four of the six study participants (4/6, 67%; participants 1, 2, 5 and 6), had a single ESBL-EC ST and the remaining two study participants (2/6, 33%; participants 3 and 4), had two ESBL-EC STs **(Table 1)**.

**Table 1.** Characteristics of the ESBL-EC recovered from each study participant.

| Participant | Number of colony picks | Species | Sequence type |
| --- | --- | --- | --- |
| 1 | 6 | *E. coli* | 131 |
| 2 | 7 | *E. coli* | 131 |
| 3 | 3 | *E. coli* | 38 |
|  | 1 | *E. coli* | 1193 |
| 4 | 2 | *E. coli* | 131 |
|  | 5 | *E. coli* | 1193 |
| 5 | 7 | *E. coli* | 349 |
| 6 | 7 | *E. coli* | 38 |

We then compared the SNV calls from multiple sc-WGS and the pe-MGS for each ESBL-EC STs. While the SNV consensus sequences for the sc-WGS and pe-MGS produced by the snippy tool had many SNV calls that were unique either to the sc-WGS or the pe-MGS with little agreement between them **(Supplementary Table 4)**, the SNV calls for the sc-WGS and the pe-MGS resulting from our custom workflow corresponded well **(Table 2, Supplementary Table 5),** with 856 697 SNVs identical by both methods, and 17 SNVs present in only one or different nucleotides. Five of the eight ESBL-EC STs (5/8, 62.5%) had identical SNV calls in the sc-WGS and pe-MGS. Three of the STs (3/10, 30%) had SNV calls unique to the sc-WGS data and which were not called in the pe-MGS data. One sample contained a ST (1/10, 10%) with SNVs unique to the pe-MGS data and that was not present in the sc-WGS data; and one ST had a SNV call that was present in both the sc-WGS and the pe-MGS data but was a different variant (1/10, 10%) **(Table 2)**.

**Table 2.**
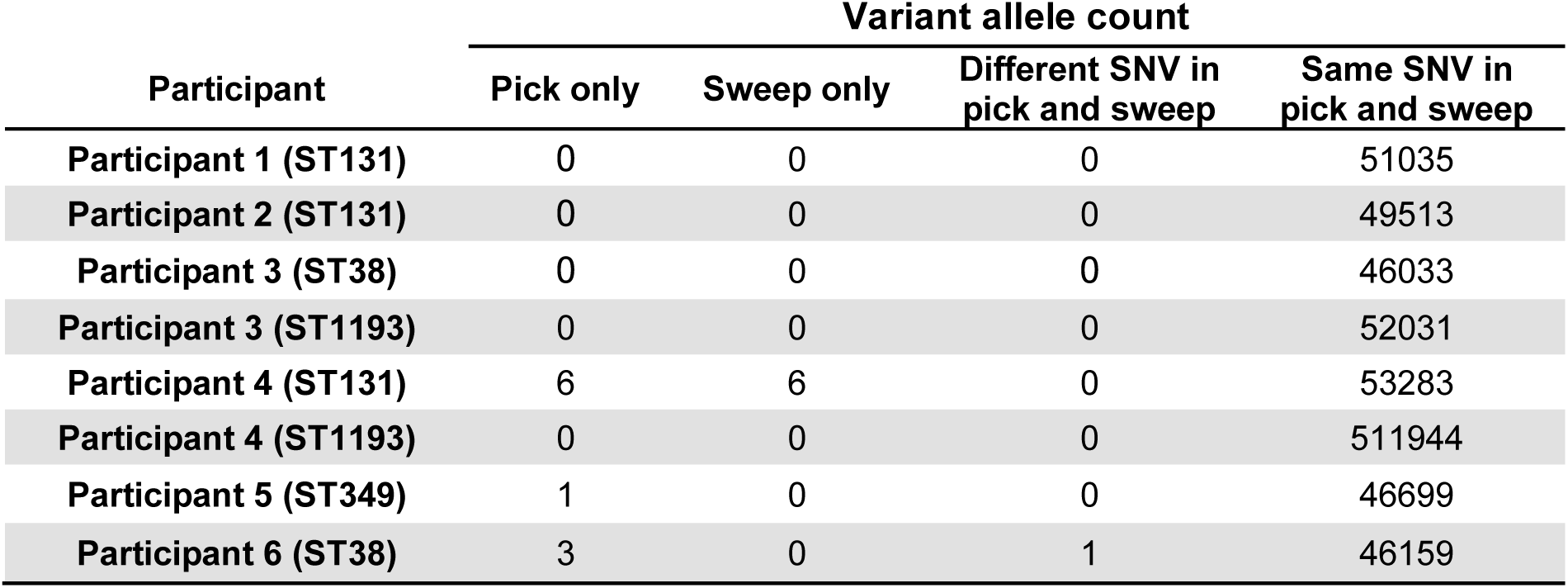
Comparison of the number of SNV calls in the sc-WGS and pe-MGS data from each study participant using the custom SNV calling workflow with the core-gene reference

### Required sequence depth for plate sweep SNV calls

We sequenced the plate sweep samples at a sequencing depth of 6Gb yielding an average of more than 1 000 x coverage for the ESBL-EC strains from each participant. To approximate the sequencing depth which will allow us to capture SNV from plates sweep with a similar accuracy to SNV calls from multiple sc-WGS, we subsampled the sequence data and fitted a logistic regression model to the resulting number of sweep SNV calls **(Supplementary Table 6**). We were able to capture 99% of all the SNV calls with 50 x sequence coverage in five ESBL-EC isolates (5/8, 62.5%), 170 x coverage in two ESBL-EC isolates (2/8, 25%) and 250 x coverage in one ESBL-EC isolate (1/8, 12.5%) **(Figure 3).**

**Figure 3.**
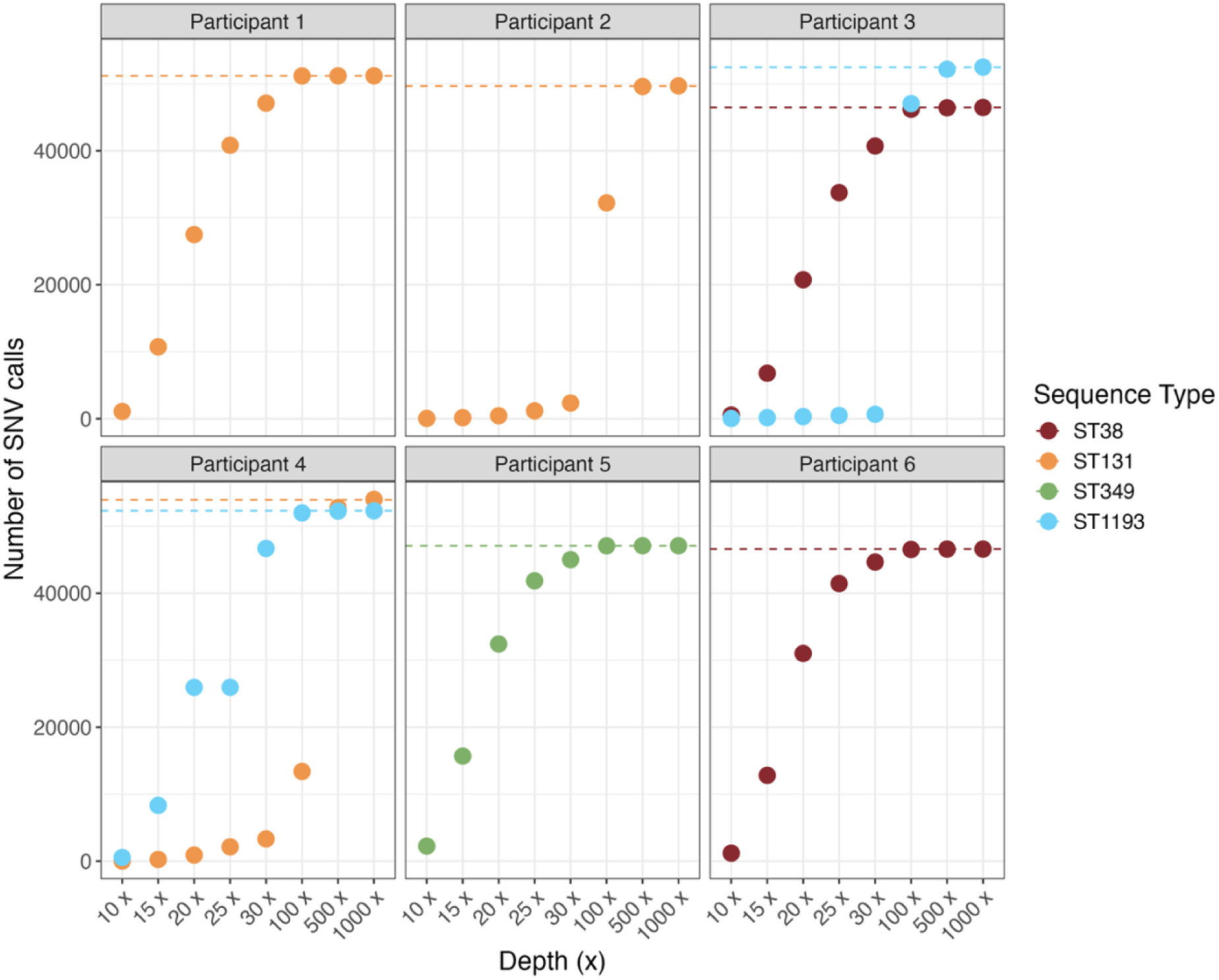
Number of SNV calls when compared to the core-gene E. coli reference at different sequence depths using subsampled sequence data from the plate sweeps for each participant. The horizontal dashed line represents the fixed combined single pick SNV calls for each lineage (ST) within each sample.

### Plate sweep method offers a cheaper, efficient alternative to multiple colony picks for describing ESBL-EC diversity

To compare the cost of each method (sc-WGS, multiple sc-WGS and pe-MGS) for the analysis of the within-host diversity of ESBL-EC, we took into consideration the cost of the microbiology method, which includes pre-enrichment in BPW, plating on selective MacConkey agar and DNA extraction, and the sequencing costs, including library preparation. The single colony pick method was the cheapest, costing around half as much as the plate sweep method **(Supplementary Table 7)**. However, when comparing the cost of multiple sc-WGS to pe-MGS, the multiple colony picks method was substantially more expensive than the plate sweep method **(Supplementary Table 7)**. Additionally, the plate sweep method required one less standard working day of microbiological processing time than single and multiple colony picks **(Supplementary Table 7)**.

## Discussion

To accurately trace the transmission of ESBL-EC, SNV-level analysis provides the highest resolution possible to date. Since analysis of a single colony is insufficient for capturing the full within-host ESBL-EC diversity, and metagenomics cannot provide sufficient resolution for *E. coli* within complex samples without expensive deep sequencing, the plate sweep method offers a more feasible alternative. Here, we validated the accuracy of plate sweep ESBL-EC SNV calls using a custom SNV calling workflow by directly comparing it to the SNV calls from seven single colony picks for six study participants. We demonstrated that by using our custom workflow with a core-gene reference, high-confidence SNV calls from plate sweep sequence data were equivalent to SNV calls in multiple single colony picks, and the approach reduced cost and time.

While pe-MGS has been used previously to describe SNV diversity in clinical samples^19^, we present a direct comparison to SNV calls from multiple sc-WGS from the same plate. Previous analyses of within-ST diversity of clinical samples have relied on data from multiple sc-WGS to make accurate SNV calls^10,47–49^, and have concluded that sequence data analysis should be able to accurately identify a difference of 10 to 17 SNVs between isolates across an *E. coli* genome^50,51^ . We demonstrate here that the pe-MGS offers a cheaper, time-efficient alternative for analysing within-ST ESBL-EC diversity.

However, the accuracy of the pe-MGS SNV calls is dependent on the reference used. Complete genome references contain hypervariable regions such as prophage regions^52^, surface polysaccharides^53^ and genomic islands^54^ that will inevitably have an increased number of associated SNVs. Accurately captured SNV variability at these sites between two isolates does not necessarily indicate two bacteria are part of distinct transmission chains. Furthermore, these regions are challenging given their often repetitive nature, providing issues to both reference assemblies and subsequent mapping, and false positive SNV calls have been observed in regions surrounding indels within the reference sequence as well as in multi-copy genes that are difficult to identify and remove with quality and depth filters^55,56^. Indeed, the use of complete genome references in our study resulted in SNV calls in our double spike sample that we expected to be clonal, i.e. no SNV calls. A possible solution would be to exclude regions of hypervariability using tools such as Dustmasker^57^. However, for our use case in the analysis of epidemiologically informative SNVs, we chose to remove any uncertainty surrounding hypervariable regions and continued with a core-gene reference that also allows for much higher reproducibility for samples with different compositions. Based on a large-scale analysis of diverse *E. coli* genomes, as it provides a reference much more equally distant to all STs regarding gene composition than a specific reference genome from a certain ST.

Additionally, we included a sliding window filter in our workflow to remove aggregated SNVs. Spatial clustering of SNVs can arise from homologous recombination, gene or genetic region (for example proteins with repetitive domain profiles) duplication, or mobile element insertion. These SNV clusters can complicate analyses and are prone to assembly errors. as repetitive sequences are challenging to resolve in particular with short reads, and subsequenctly are also prone to misinterpretation of transmission, thus we chose to remove them using a sliding window approach^18^.

An accurate analysis of the within-host diversity using pe-MGS has been used successfully for identification of transmission routes^18,30^. While heterozygous SNVs are yet not commonly considered when analysing bacterial phylogenetic relationships^19,30^, inclusion of heterozygous SNVs has proven invaluable in transmission inference^18^. Indeed, the incorporation of SNV data into algorithms have provided accurate insights into the transmission rates in large cohort datasets^37^. Here, we observed that with the inclusion of heterozygous SNVs in the pe-MGS data analysis, the resulting within-host SNV calls closely resembled the collective SNV calls from multiple sc-WGS, reiterating the power of pe-MGS data.

While we have fully validated a workflow for SNV calling using pe-MGS data from a plate sweep, this study was limited to six rectal swab samples with a maximum of two ESBL-EC STs in two of the samples. Therefore, we do not know the SNV call accuracy, or the coverage required to accurately call SNVs in samples with three or more STs, which will need to be validated in the future. The use of a core-gene list as a reference is also limiting since it is not a fully assembled genome, leading to breaks in mapping at gene boundaries, and requires a large collection of genomes to construct. More broadly, the pe-MGS method we assessed uses short-read sequencing and would not be expected to be able to recover mobile genetic elements like plasmids well, nor associate them with bacterial strains, thus is currently most useful for epidemiological purposes.

## Conclusion

Consideration of within-host diversity of ESBL-EC present in a given sample allows for a more complete picture of colonisation at a certain time point. If the full diversity of ESBL-EC within the stool sample is not considered, some isolates may be misclassified as being acquired when they were present throughout the sampling period at low levels^50^. The custom SNV calling workflow we have developed here can provide a cost-effective method for defining within-host diversity at scale which will allow more accurate description of transmission events. This in turn will allow us to better understand transmission patterns of particular pathogens, and to design, test and optimise infection prevention and control strategies to block transmission.

## Ethics

The collection of stool from healthy volunteers was collected with written consent for the use in this study (LSTM Research Tissue Bank RTB/2022/007). The collection of clinical samples for the TRACS-Liverpool study was approved by the National Research Ethics Service Greater Manchester South ethics committee (ref: 22/NW/0343). Written informed consent was obtained from participants or consultees, as appropriate.

## Data Availability

All data produced in the present work are contained in the manuscript.

https://www.ebi.ac.uk/ena/browser/view/PRJEB101999

## Acknowledgements

We would like to thank Ross Gray for the illustrations used in graphical abstract of this manuscript as well as all the clinical staff, study participants and volunteers who facilitated this study. The authors also wish to acknowledge CSC – IT Center for Science, Finland, for computational resources.

## Author contributions

Conceptualization and method development were done by S.G., E.H., N.A.F., F.E.G. and J.M.L. Investigation was undertaken by S.G., S.F., E.P.-B, C.M., and M.M. Methodology was done by S.G., T.M. and J.M.L. Data analysis was done by S.G., T.M. and J.M.L. The original draft was prepared by S.G., E.H., T.M., J.C., N.A.F., F.E.G and J.M.L. and then reviewed and edited by all authors. Supervision was provided by E.H., N.A.F., F.E.G. and J.M.L.

## Reference

1. Klepac, P., Funk, S., Hollingsworth, T. D., Metcalf, C. J. E. & Hampson, K. Six challenges in the eradication of infectious diseases. Epidemics 10, 97–101 (2015).

2. Balloux, F. et al. From Theory to Practice: Translating Whole-Genome Sequencing (WGS) into the Clinic. Trends Microbiol 26, (2018).

3. Jiang, Y., Jiang, Y., Wang, S., Zhang, Q. & Ding, X. Optimal sequencing depth design for whole genome re-sequencing in pigs. BMC Bioinformatics 20, 556 (2019).

4. Martischang, R. et al. Epidemiology of ESBL-producing Escherichia coli from repeated prevalence studies over 11 years in a long-term-care facility. Antimicrob Resist Infect Control 10, 148 (2021).

5. Raffelsberger, N. et al. Community carriage of ESBL-producing Escherichia coli and Klebsiella pneumoniae: a cross-sectional study of risk factors and comparative genomics of carriage and clinical isolates. mSphere 8, e00025–23 (2023).

6. Doughty, E. L. et al. Endemicity and diversification of carbapenem-resistant Acinetobacter baumannii in an intensive care unit. Lancet Reg Health West Pac 37, (2023).

7. Tong, S. Y. C. et al. Genome sequencing defines phylogeny and spread of methicillin-resistant Staphylococcus aureus in a high transmission setting. Genome Res 25, 111–118 (2015).

8. Msefula, C. L. et al. Genotypic Homogeneity of Multidrug Resistant S. Typhimurium Infecting Distinct Adult and Childhood Susceptibility Groups in Blantyre, Malawi. PLoS One 7, e42085 (2012).

9. Lidin-Janson, G., Kaijser, B., Lincoln, K., Olling, S. & Wedel, H. The homogeneity of the faecal coliform flora of normal school-girls, characterized by serological and biochemical properties. Med Microbiol Immunol 164, 247–253 (1978).

10. Schlager, T. A., Hendley, J. O., Bell, A. L. & Whittam, T. S. Clonal diversity of Escherichia coli colonizing stools and urinary tracts of young girls. Infect Immun 70, 1225–1229 (2002).

11. Yang, A. R., Bowling, J. M., Morgan, C. E., Bartram, J. & Kayser, G. L. Predictors of household drinking water E. coli contamination: Population-based results from rural areas of Ghana, Malawi, Mozambique, Niger, Rwanda, Uganda, and Zambia. Int J Hyg Environ Health 264, 114507 (2025).

12. Quince, C., Walker, A. W., Simpson, J. T., Loman, N. J. & Segata, N. Shotgun metagenomics, from sampling to analysis. Nat Biotechnol 35, 833–844 (2017).

13. Oechslin, C. P. et al. Limited Correlation of Shotgun Metagenomics Following Host Depletion and Routine Diagnostics for Viruses and Bacteria in Low Concentrated Surrogate and Clinical Samples. Front Cell Infect Microbiol 8, 403892 (2018).

14. Foster-Nyarko, E. & Pallen, M. J. The microbial ecology of Escherichia coli in the vertebrate gut. FEMS Microbiol Rev 46, fuac008 (2022).

15. Mäklin, T. et al. Bacterial genomic epidemiology with mixed samples. Microb Genom 7, 000691 (2021).

16. Maklin, T. et al. High-resolution sweep metagenomics using fast probabilistic inference [version 1; peer review: 1 approved, 1 approved with reservations]. Wellcome Open Res 5, 1–20 (2020).

17. Lewis, J. M. et al. Colonization dynamics of extended-spectrum beta-lactamase-producing Enterobacterales in the gut of Malawian adults. Nat Microbiol 7, 1593–1604 (2022).

18. Tonkin-Hill, G. et al. Pneumococcal within-host diversity during colonization, transmission and treatment. Nature Microbiology 2022 7:*11* 7, 1791–1804 (2022).

19. Thorpe, H. A. et al. Pan-pathogen deep sequencing of nosocomial bacterial pathogens in Italy in spring 2020: a prospective cohort study. Lancet Microbe 100890 (2024) doi:10.1016/S2666-5247(24)00113-7.

20. Sibale, L. L. et al. Within-host genetic diversity of pneumococcal serotype 3 during one-year prolonged carriage in a healthy adult. Nature Communications 2025 16:1 16, 1–12 (2025).

21. Schwaber, M. J. & Carmeli, Y. Mortality and delay in effective therapy associated with extended-spectrum beta-lactamase production in Enterobacteriaceae bacteraemia: a systematic review and meta-analysis. J Antimicrob Chemother 60, 913–920 (2007).

22. Melzer, M. & Petersen, I. Mortality following bacteraemic infection caused by extended spectrum beta-lactamase (ESBL) producing E. coli compared to non-ESBL producing E. coli. Journal of Infection 55, 254–259 (2007).

23. Blom, A., Ahl, J., Månsson, F., Resman, F. & Tham, J. The prevalence of ESBL-producing Enterobacteriaceae in a nursing home setting compared with elderly living at home: a cross-sectional comparison. 10.1186/s12879-016-1430-5(2016) doi:10.1186/s12879-016-1430-5.

24. TRACS Liverpool Part 2 - Health Research Authority. https://www.hra.nhs.uk/planning-and-improving-research/application-summaries/research-summaries/tracs-liverpool-part-2/.

25. Gallichan, S. et al. Optimized methods for the targeted surveillance of extended-spectrum beta-lactamase-producing Escherichia coli in human stool. Microbiol Spectr 13, (2025).

26. ThermoFisher Scientific. User Guide: Qubit dsDNA HS Assay Kits. https://tools.thermofisher.com/content/sfs/manuals/Qubit_dsDNA_HS_Assay_UG.pdf.

27. 27. Agilent Technologies. Agilent 4150 TapeStation System Manual. https://www.agilent.com/cs/library/usermanuals/public/4150-TapeStation_SystemManual.pdf.

28. Chen, S., Zhou, Y., Chen, Y. & Gu, J. fastp: an ultra-fast all-in-one FASTQ preprocessor. Bioinformatics 34, i884–i890 (2018).

29. Andrews, S. FastQC: A Quality Control tool for High Throughput Sequence Data. https://www.bioinformatics.babraham.ac.uk/projects/fastqc/ (2010).

30. Khawaja, T. et al. Deep sequencing of Escherichia coli exposes colonisation diversity and impact of antibiotics in Punjab, Pakistan. Nature Communications 2024 15:*1* 15, 1–11 (2024).

31. Lees, J. A. et al. Fast and flexible bacterial genomic epidemiology with PopPUNK. Genome Res 29, 304–316 (2019).

32. Mäklin, T. Escherichia coli lineage deconvolution indexes for Themisto, mSWEEP/mGEMS, and demix_check. 10.5281/ZENODO.12528310 doi:10.5281/ZENODO.12528310.

33. Alanko, J. N., Vuohtoniemi, J., Mäklin, T. & Puglisi, S. J. Themisto: a scalable colored k-mer index for sensitive pseudoalignment against hundreds of thousands of bacterial genomes. bioRxiv 2023.02.24.529942 (2023) doi:10.1101/2023.02.24.529942.

34. harry-thorpe/demix_check. https://github.com/harry-thorpe/demix_check.

35. Ondov, B. D. et al. Mash: fast genome and metagenome distance estimation using MinHash. Genome Biol 17, (2016).

36. Hunt, M. et al. ARIBA: rapid antimicrobial resistance genotyping directly from sequencing reads. Microb Genom 3, (2017).

37. Tonkin-Hill, G. et al. Enhanced metagenomics-enabled transmission inference with TRACS. *bioRxiv* 2024.08.19.608527 (2024) doi:10.1101/2024.08.19.608527.

38. Wirth, T. et al. Sex and virulence in Escherichia coli: an evolutionary perspective. Mol Microbiol 60, 1136–1151 (2006).

39. Horesh, G. et al. A comprehensive and high-quality collection of escherichia coli genomes and their genes. Microb Genom 7, 1–15 (2021).

40. Shen, W., Le, S., Li, Y. & Hu, F. SeqKit: A Cross-Platform and Ultrafast Toolkit for FASTA/Q File Manipulation. PLoS One 11, e0163962 (2016).

41. Garrison, E. & Marth, G. Haplotype-based variant detection from short-read sequencing. https://arxiv.org/abs/1207.3907v2 (2012).

42. Danecek, P. et al. Twelve years of SAMtools and BCFtools. Gigascience 10, 1–4 (2021).

43. Köster, J. et al. Sustainable data analysis with Snakemake. F1000Research *2021 10:33* 10, 33 (2021).

44. lh3/seqtk: Toolkit for processing sequences in FASTA/Q formats. https://github.com/lh3/seqtk.

45. Baty, F. et al. A Toolbox for Nonlinear Regression in R: The Package nlstools. J Stat Softw 66, 1–21 (2015).

46. Pokharel, P., Dhakal, S. & Dozois, C. M. The Diversity of Escherichia coli Pathotypes and Vaccination Strategies against This Versatile Bacterial Pathogen. Microorganisms 2023, Vol. 11, *Page 344* **11**, 344 (2023).

47. Pallen, M. J. et al. Genomic diversity of Escherichia coli from healthy children in rural Gambia. 10.7717/peerj.10572 doi:10.7717/peerj.10572.

48. Dixit, O. V. A., O’Brien, C. L., Pavli, P. & Gordon, D. M. Within-host evolution versus immigration as a determinant of Escherichia coli diversity in the human gastrointestinal tract. Environ Microbiol 20, 993–1001 (2018).

49. Ludden, C. et al. Defining nosocomial transmission of Escherichia coli and antimicrobial resistance genes: a genomic surveillance study. Lancet Microbe 2, e472–e480 (2021).

50. Ludden, C. et al. Defining nosocomial transmission of Escherichia coli and antimicrobial resistance genes: a genomic surveillance study. Lancet Microbe 2, e472–e480 (2021).

51. Nguyen, M. N. et al. Tracing carriage, acquisition, and transmission of ESBL-producing Escherichia coli over two years in a tertiary care hospital. Genome Med 16, 1–15 (2024).

52. Huey, B. & Hall, J. Hypervariable DNA fingerprinting in Escherichia coli: minisatellite probe from bacteriophage M13. J Bacteriol 171, 2528 (1989).

53. Liu, B. et al. Structure and genetics of Escherichia coli O antigens. FEMS Microbiol Rev 44, 655 (2019).

54. Moritz, R. L. & Welch, R. A. The Escherichia coli argW-dsdCXA Genetic Island Is Highly Variable, and E. coli K1 Strains Commonly Possess Two Copies of dsdCXA. J Clin Microbiol 44, 4038 (2006).

55. Bush, S. J. Generalizable characteristics of false-positive bacterial variant calls. Microb Genom 7, 000615 (2021).

56. Li, H. & Wren, J. Toward better understanding of artifacts in variant calling from high-coverage samples. Bioinformatics 30, 2843–2851 (2014).

57. Morgulis, A., Gertz, E. M., Schäffer, A. A. & Agarwala, R. A fast and symmetric DUST implementation to mask low-complexity DNA sequences. J Comput Biol 13, 1028–1040 (2006).

